# Lifestyle behaviours of children and adolescents during the first two waves of the COVID-19 pandemic in Switzerland and their relation to well-being: a population-based study

**DOI:** 10.1101/2021.11.29.21267019

**Authors:** Gabriela P. Peralta, Anne-Linda Camerini, Sarah R. Haile, Christian R. Kahlert, Elsa Lorthe, Laura Marciano, Andres Nussbaumer, Thomas Radtke, Agne Ulyte, Milo A. Puhan, Susi Kriemler

**Author notes:** **Corresponding author:** Prof. Dr. med. Susi Kriemler. GPP and ALC contributed equally and share first authorship. MAP and SK contributed equally and share last authorship.

## Abstract

**Background:** Previous studies assessing the impact of the COVID-19 pandemic on children’s and adolescent’s lifestyle focused mainly on the first wave in early 2020. We aimed to describe changes in adherence to recommendations for physical activity (PA), screen time (ST), and sleep duration over the first two waves of the pandemic (March-May 2020 and October 2020-January 2021) in Switzerland, and to assess the associations of these lifestyle behaviours with life satisfaction and overall health, as indicators of well-being.

**Methods:** We included 3168 participants aged 5 to 18 years from four Swiss cantons. Participants or their parents completed repeated questionnaires and reported on their (child’s) PA, ST, sleep, life satisfaction, and overall health. We analysed lifestyle behaviours in terms of adherence to international recommendations. We used linear and logistic regression models to assess the associations of number of recommendations met and adherence patterns with well-being indicators.

**Findings:** Compared to the pre-pandemic period, the percentage of participants meeting the recommendations for PA and ST decreased strikingly during March-May 2020, while there was a slight increase in those meeting recommendations for sleep. During October 2020-January 2021, the percentage of compliant children for PA and ST increased but remained lower than before the pandemic. Participants meeting all three recommendations were more likely to report excellent health (OR: 1·87 [1·15-3·08]) and a higher life satisfaction score (β: 0·59 [0·30-0·88]) than participants not meeting any recommendation. Adherence to recommendations for PA and sleep, PA and ST, and sleep and ST was similarly associated with both well-being indicators.

**Interpretation:** We show a substantial impact of the COVID-19 pandemic on children’s and adolescents’ lifestyle behaviours with a partial recovery over time, and an association between lifestyle and well-being. Public health policies to promote children’s and adolescents’ well-being should target PA, ST, and sleep simultaneously.

**Funding:** Corona Immunitas.

**RESEARCH IN CONTEXT:** *Evidence before the study:* We searched PubMed for studies assessing the effects of the COVID-19 pandemic on children’s and adolescents’ lifestyle behaviours, published up to September 6, 2021, with no language restrictions. Of the studies found, nearly all compared lifestyle behaviours before and during the strict confinement in the first wave of the pandemic, and very few studies extended their assessment beyond June 2020. The only longitudinal study assessing lifestyle changes up to 2021 included a sample of nineteen boys. Some studies assessed the association between lifestyle behaviours and well-being after the outbreak of the pandemic, but all used a single-behaviour approach (i.e., evaluated only one lifestyle behaviour) and no study considered the combined contribution of physical activity (PA), screen time (ST), and sleep. In addition, most studies were cross-sectional and did not consider pre-pandemic lifestyle behaviours.

*Added value of this study:* This is the first study assessing changes in adherence to international recommendations regarding PA, ST, and sleep duration in children and adolescents during the first two waves of the COVID-19 pandemic and the joint associations of these lifestyle behaviours with well-being. We used data from 3168 children and adolescents from four different Swiss cantons. We included measurements of PA, ST, and sleep before the pandemic, during the first wave between March and May 2020, and during the second wave between October 2020 and January 2021. We also assessed life satisfaction and overall health as indicators of well-being, between January and April 2021. We showed that, compared to the pre-pandemic period, the proportion of children and adolescents following the recommendations for PA and ST decreased during the first wave (lockdown period), while there was a slight increase in those meeting recommendations for sleep. During the second wave, the prevalence of compliant children and adolescents for PA and ST recovered but remained below pre-pandemic levels. Furthermore, we found an association between the number of recommendations met for lifestyle behaviours during the second wave and well-being assessed between January and April 2021. In contrast, there was no such association for the periods before the pandemic or during the lockdown. Participants following the recommendations for all three lifestyle behaviours or for combinations of two of them in the second wave were more likely to report excellent health and had a higher life satisfaction score, than those not meeting any recommendation.

*Implications of all the available evidence:* Our findings demonstrate that the COVID-19 pandemic has had a strong negative effect on children’s and adolescents’ lifestyle behaviours, but some recovery has taken place within the first year since the outbreak. Policymakers should imperatively consider the balance of disease prevention and promotion of a healthy lifestyle when (re-)activating restrictive measures. Given the already high prevalence of children and adolescents not meeting lifestyle recommendations in the pre-pandemic period, our findings highlight the urgent need for public health policies aiming to avoid permanent negative changes on children’s and adolescents’ lifestyle and to mitigate the health risks associated with adverse changes during the pandemic. In addition, our study indicates that lifestyle is an important predictor of children’s and adolescents’ well-being, and it further suggests that future public health strategies aiming to promote well-being should target sufficient time for PA and sleep as well as reduce ST.

## INTRODUCTION

The COVID-19 pandemic and its associated confinement measures have led to significant changes in the daily life of children and adolescents. Like many countries, the Swiss government imposed a strict lockdown between March and May 2020.^1^ During that period, classroom teaching was interrupted, and then continued with a partial return to class (i.e., alternating face-to-face and online classes) until the start of the summer break. In August 2020, schools reopened as normal, though with strict prevention measures in place. In autumn and early winter 2020, Switzerland experienced an alarming increase of SARS-CoV-2 infections. Although classroom teaching in primary and secondary education was secured, sports clubs, gyms, and other leisure institutions were closed to some extent (i.e., open only for children under age 16 years) or had restrictions in place until early 2021.^2^

Previous research reported that COVID-19 restrictions during the first semester of 2020 resulted in a drastic reduction of physical activity (PA) and increase in screen time (ST) among children and adolescents.^3^ A study conducted in Shanghai, showed that the prevalence of physically inactive students (aged 6 to 17 years) increased from 21% to 66% during the first months of the pandemic.^4^ Similarly, other studies in Germany and Canada reported a substantial decrease in the prevalence of children and adolescents meeting the recommendations of up to two hours per day for ST^5^ during the lockdown in early 2020.^6,7^ In addition, restrictions led to a delay of sleep onset and offset resulting in a subtle increase of sleep duration during that period.^3^ Considering the pre-existing evidence of an association between lifestyle behaviours and well-being in children and adolescents,^8,9^ some studies examined this association during the pandemic. They reported that higher levels of PA were associated with better well-being, while higher levels of ST were related to decreased emotional well-being.^10–12^

Despite the existing literature, further research is needed for a more comprehensive understanding of the effects of the COVID-19 pandemic on children’s and adolescents’ lifestyle, and on the relationship between lifestyle behaviours and well-being. Very few studies extended their assessment beyond June 2020,^12–14^ and only one small (n=19) longitudinal study assessed lifestyle changes up to 2021.^15^ In addition, studies assessing the associations between lifestyle and well-being during the pandemic were all based on a single-behaviour approach. Nevertheless, there is evidence supporting that meeting recommendations for more than one lifestyle behaviour is more beneficial for children’s and adolescents’ health and development than meeting the recommendations for a single behaviour.^16,17^ A recent study using data from before the pandemic reported that a combination of lower ST and higher PA appears to have joint and positive associations with mental well-being in adolescents.^18^ However, the study could not account for the contribution of sleep duration to this association. This highlights the need for further research including repeated measurements of lifestyle beyond the early phases of the pandemic and assessing the combined associations of lifestyle behaviours and well-being.

Therefore, we aimed to describe changes in adherence to international recommendations for PA, ST, and sleep duration over the first two waves of the COVID-19 pandemic (March-May 2020 and October 2020-January 2021) in children and adolescents aged 5 to 18 years from four Swiss cantons, and to assess the combined associations of these lifestyle behaviours with overall health and life satisfaction.

## METHODS

### Study design and participants

This population-based observational study is part of *Corona Immunitas*, a research network that investigates the spread and impact of COVID-19 pandemic in Switzerland.^19^ We included children and adolescents from four (out of 26) Swiss cantons: Ticino (TI), St. Gallen (SG), Graubünden (GR), and Zurich (ZH), which include 30% of the Swiss population. In TI, SG, and GR, participants were recruited based on a representative sample of children and adolescents drawn from the Swiss Federal Registry. In ZH, the data come from the *Ciao Corona* study,^20^ a cohort of school-aged children from 55 schools. Primary schools were randomly selected from the list of all schools in the canton of ZH and matched with the geographically closest secondary school.

In this study, we included participants aged 5 to 18 years with baseline data collected between June 2020 and January 2021 and follow-up data collected between January and April 2021. In ZH, parents were asked to complete the questionnaires together with their child. In the other three cantons, parents completed the questionnaires for children aged 5 to13 years, while adolescents aged 14 to 18 years completed the questionnaires on their own. The study was approved by the Ethics Committee of all regions (TI: 2020-01514; SG/GR: 2020-01247; ZH: 2020-01336). All participants and/or their parents provided informed consent. See further details on participant recruitment and data collection in the appendix.

### Measures

#### Lifestyle

We assessed PA, ST, and sleep with reference to the period before the pandemic (i.e., before March 2020) and during the lockdown in the first wave (i.e., between 16 March and 10 May 2020) in the baseline questionnaire. Lifestyle during the second wave (i.e., between October 2020 and January 2021) was also assessed in the baseline questionnaire in TI, SG and GR, while in ZH it was assessed in a separate follow-up questionnaire (Table S1).

PA was assessed by asking how many hours, on average, participants spent with PA (at least with light sweating) per week. Weekly hours were converted to daily hours. ST was assessed by asking how many hours, on average, participants spent with electronic media (e.g., smartphone, computer, PlayStation, Xbox, Nintendo, TV) on a typical weekday and on a typical weekend day. Sleep was assessed by asking how many hours, on average, participants slept on a typical weekday and on a typical weekend day. For ST and sleep, we calculated a weighted average as follows: [(weekday*5) + (weekend day*2)]/7. In ZH, there were some variations in the formulation of the questions and the calculation of daily hours (Table S2).

We analysed lifestyle in terms of adherence to international recommendations: ≥ 1h/day of PA,^21^ ≤2h/day of ST,^5^ and age-specific sleep duration (10-13 h/night for 5 years; 9-11 h/night for 6 to 13 years; 8-10 h/night for 14 to 17 years; 7-9 h/night for 18 years).^22^ We also classified participants into one of the following recommendation adherence patterns: none, PA only, sleep only, ST only, PA and sleep, PA and ST, sleep and ST, and all three.

#### Well-being

We measured current life satisfaction and overall health as indicators of well-being in a follow-up questionnaire between January and April 2021. This was the first follow-up questionnaire after baseline in TI, SG and GR and the second follow-up questionnaire after baseline in ZH. Well-being was thus measured two to three months after the assessment of lifestyle behaviours with reference to the second wave (Table S1).

We assessed life satisfaction with the Cantril ladder, a visual scale for rating how children and adolescents perceive their life on a 11-point scale ranging from ‘0-the worst possible life’ to ‘10-the best possible life’. We assessed overall health by asking ‘How would you rate your/your child’s health?’. Respondents were asked to describe their (child’s) health status on a scale from ‘1-poor’ to ‘4-excellent’.

Both items have been widely used in epidemiological research.^23,24^ We analysed life satisfaction as a continuous variable and overall health as a binary variable (‘excellent’ versus lower ratings).

#### Covariates

We assessed participants’ sex, age, height, and weight in the baseline questionnaire. We also collected information on parents’ nationality (at least one Swiss; both non-Swiss) and educational level (high: at least one with preparatory high school to university; low/medium: both up to apprenticeship or professional school). We used height and weight to derive age- and sex-specific body mass index (BMI) z-scores by using the WHO Child Growth Standards, and categorised it as underweight, normal weight, and overweight according to the guidelines.^25^ We categorised participants’ age as follows: 5 to 9 years, 10 to 14 years, and 15 to 18 years.

### Statistical analysis

We conducted descriptive analysis stratified by age category. To assess the associations of the number of recommendations met and the adherence patterns with overall health and life satisfaction, we used multivariable logistic and linear regression, respectively. We adjusted the models for child’s sex, age category, BMI category, parents’ nationality, educational level, and canton. We first tested a model including only the number of recommendations met/adherence patterns before the pandemic and the covariates. Then, we added the lifestyle measures for the lockdown period and, finally, for the second wave.

We used a complete case strategy for the multivariate analysis and reported missing data in the Table 1 footnotes. A further analysis, imputing well-being indicators and covariates, yielded very similar results (data not shown). We performed the statistical analysis in R (version 4.0.3). See further details on statistical analysis in the appendix.

**Table 1.** Characteristics of the study sample*.

| **Characteristics** | **Overall**<br>n= 3168 | **ZH**<br>n= 1887 | **TI**<br>n= 886 | **SG**<br>n=198 | **GR**<br>n=197 |
| --- | --- | --- | --- | --- | --- |
| **Sex. Girls** | 1651 (52.1) | 982 (52.0) | 448 (50.6) | 108 (54.5) | 113 (57.4) |
| **Age** |  |  |  |  |  |
| 5-9 y | 1006 (31.8) | 602 (31.9) | 268 (30.2) | 68 (34.3) | 68 (34.5) |
| 10-14 y | 1734 (54.7) | 1219 (64.6) | 337 (38.0) | 88 (44.4) | 90 (45.7) |
| 15-18 y | 428 (13.5) | 66 (3.5) | 281 (31.7) | 42 (21.2) | 39 (19.8) |
| **BMI** |  |  |  |  |  |
| Underweight | 138 (4.4) | 85 (4.5) | 44 (5.1) | 1 (0.5) | 8 (4.3) |
| Normal weight | 2487 (79.7) | 1487 (79.2) | 682 (78.5) | 154 (83.2) | 164 (87.7) |
| Overweight | 494 (15.8) | 306 (16.3) | 143 (16.5) | 30 (16.2) | 15 (8.0) |
| **Parental nationality** |  |  |  |  |  |
| Swiss | 2724 (86.3) | 1613 (85.9) | 756 (85.3) | 177 (89.4) | 178 (90.4) |
| Non-Swiss | 434 (13.7) | 264 (14.1) | 130 (14.7) | 21 (10.6) | 19 (9.6) |
| **Parental education** |  |  |  |  |  |
| Low/Medium | 837 (27.1) | 427 (23.0) | 235 (27.9) | 90 (46.4) | 85 (45.0) |
| High | 2247 (72.9) | 1432 (77.0) | 607 (72.1) | 104 (53.6) | 104 (55.0) |
| **Data on well-being between January and April 2021** | 2634 (83.1) | 1650 (87.4) | 730 (82.4) | 116 (58.6) | 138 (70.1) |

Abbreviations: BMI: body mass index; GR: Graubünden; SG: St. Gallen; TI: Ticino; y: years; ZH: Zurich.
\* Data are n (%). Some percentages do not add up to 100% because of rounding. Some data were missing for BMI (49, 1.5%), parental nationality (10, 0.3%) and parental education (84, 2.7%). There were no missing data for sex and age.

## RESULTS

### Description of the study sample

We included 3168 children and adolescents in the descriptive analysis of lifestyle behaviours over time and 2634 in the regression analysis including lifestyle behaviours and well-being (Figure S1). Compared to the study samples, children and adolescents excluded from the analyses were more likely to have both parents of non-Swiss origin and with low/medium education level (Tables S3 and S4). Table 1 shows the main characteristics of the participants. Most participants had a normal BMI (80%), at least one Swiss parent (86%) and at least one parent with high education level (73%).

In the follow-up assessment between January and April 2021, 42% of participants reported excellent health. The median life satisfaction score was 8·0 (P25-P75: 7·0-9·0). Older adolescents (15 to 18 years) were less likely to report excellent health and had a lower median life satisfaction than younger participants (Table 2).

**Table 2.** Description of participants’ well-being measures between January and April 2021 by age group.

|  | **5-9 y** | **10-14 y** | **15-18 y** | **Total** |
| --- | --- | --- | --- | --- |
| **Excellent health, n (%)** | 437 (49.8) | 580 (40.3) | 101 (32.3) | 1118 (42.5) |
| **Life satisfaction\*, median (P<sub>25</sub> - P<sub>75</sub>)** | 8.0 (8.0 - 9.0) | 8.0 (7.0 - 9.0) | 7.0 (6.0 - 8.0) | 8.0 (7.0 - 9.0) |

Abbreviation: P<sub>25</sub>: percentile 25<sup>th</sup>; P<sub>75</sub>: percentile 75<sup>th</sup>; y: years.
n= 2,630 for overall health and n=2,628 for life satisfaction. \*Range: 0-worst possible life to 10-best possible life

### Changes in PA, ST, sleep, and number of recommendations met

Across all age groups, we observed a decrease in the percentage of participants meeting the individual recommendations for PA and ST between the pre-pandemic period and the lockdown period (Figure 1 and Table S4 in the appendix). While these percentages improved during the second wave, they remained below the pre-pandemic levels among the 10- to 14-year-olds and the 15- to 18-year-olds. In the 5- to 9-years-olds, PA levels, too, remained below the pre-pandemic level, but the percentage of children meeting the recommendations for ST returned to levels observed for the period before COVID-19. Adherence to recommendations for sleep also changed over time but the pattern was less consistent among age groups. Among the 10- to 14-year-olds and the 15- to 18-year-olds, the percentage of compliant participants increased slightly during the lockdown, but during the second wave it was lower than before the pandemic. For the 5- to 9-years-olds, the percentage of compliant children remained stable.

**Figure 1.**
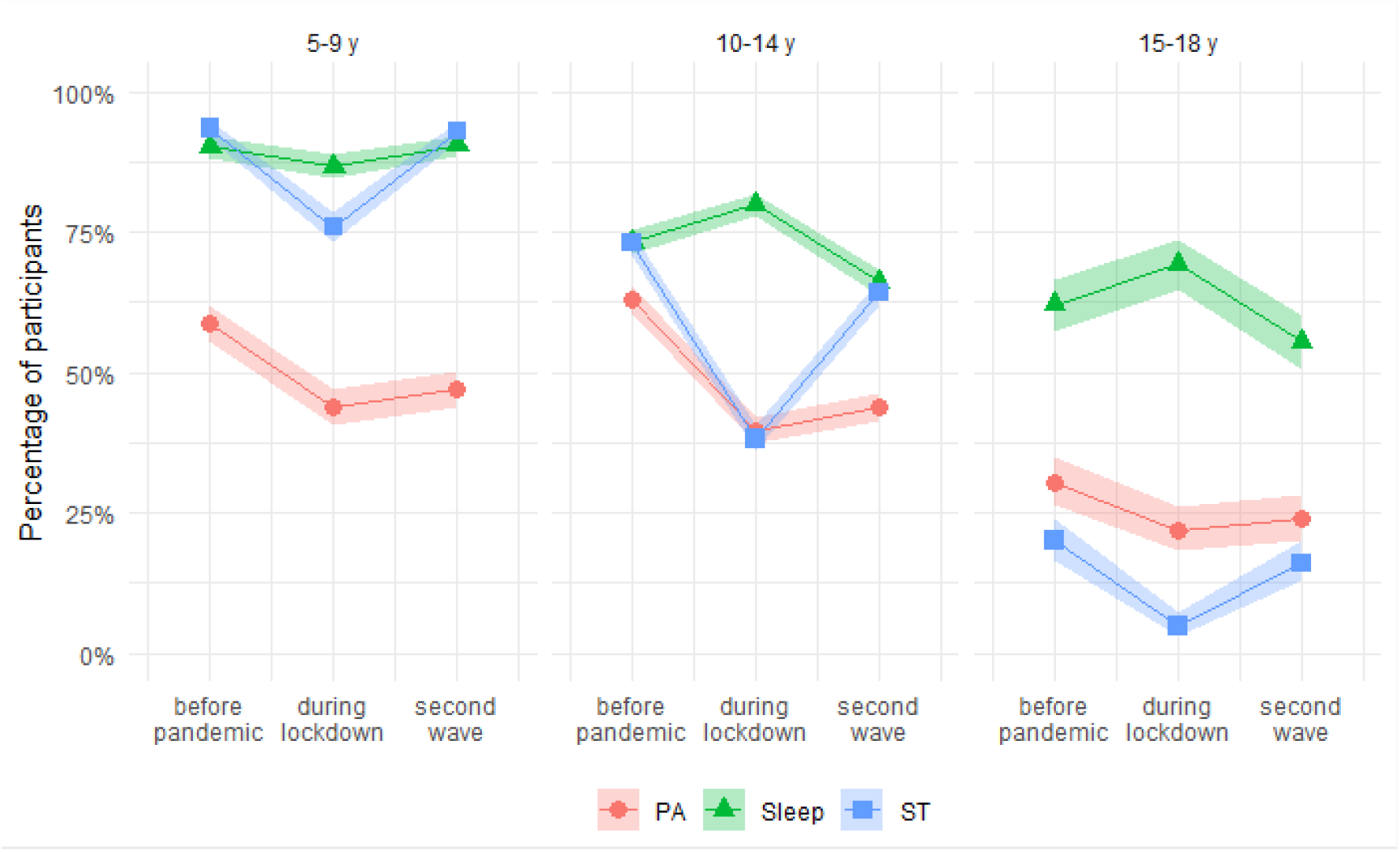
Prevalence of participants meeting recommendations for PA, ST and sleep duration by age group. Abbreviations: PA: physical activity; sleep: sleep duration; ST: screen time; y: years. Time points: before pandemic: before March 2020; during lockdown: between 16 March and 10 May 2020; second wave: between October 2020 and January 2021. Shaded area represents 95% confidence intervals. We classified participants as meeting recommendations according to international guidelines: ≥ 1h/day of PA, ≤2h/day of ST, and the recommended range by age-groups for sleep duration (i.e., 10-13 h/nigh of sleep for age 5 years, 9-11 h/night for ages 6-13 years, 8-10 h/night for ages 14-17, 7-9 h/night for age 18 years).

The percentage of participants meeting all three recommendations decreased during the lockdown period and increased again during the second wave (Figure 2 and Table S5). Yet, it remained below the percentages observed for the pre-pandemic period. At all-time points, the 15- to 18-year-olds reported the highest percentage of participants not meeting any of the recommendations.

**Figure 2.**
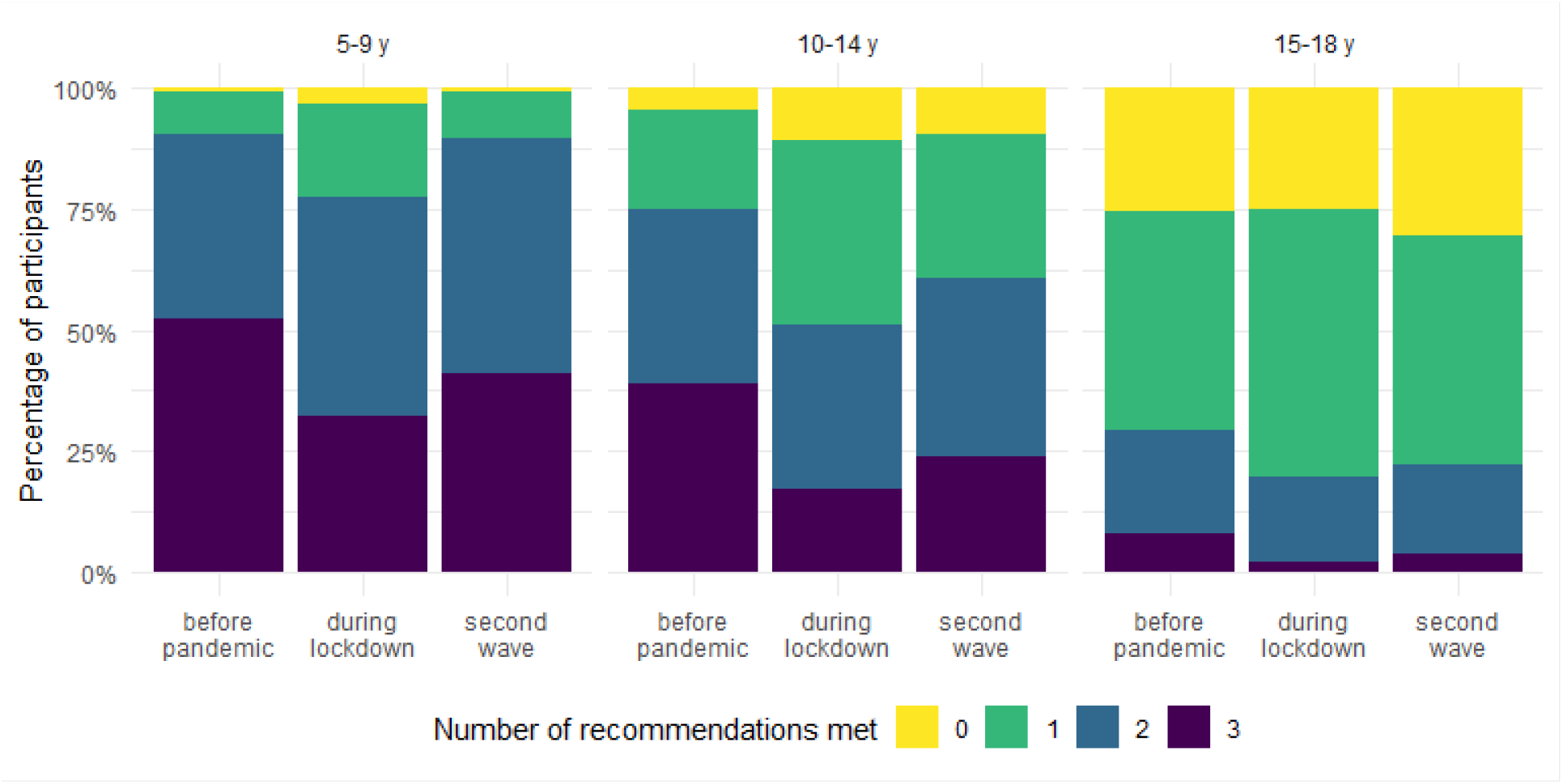
Number of recommendations met by age group. Abbreviations: y: years Time points: before pandemic: before March 2020; during lockdown: between 16 March and 10 May 2020; second wave: between October 2020 and January 2021.

We observed a similar pattern of change for boys and girls (Figures S2 and S3). For all age groups and time points, girls were less likely to meet PA recommendations. Furthermore, girls aged 15 to 18 years tended to be less compliant with ST recommendations. We also analysed lifestyle behaviours as continuous variables and observed a similar trend in changes as described when categorising the variables in terms of adherence to the recommendations (Tables S7 and S8).

### Associations of number of recommendations met and adherence patterns with well-being

Adjusted models showed that the number of recommendations met, and adherence patterns measured with reference to the second wave were stronger predictors of well-being between January and April 2021 than the same measures before the pandemic and during the lockdown (Tables 3 and 4). We found that participants meeting recommendations for all three lifestyle behaviours during the second wave were almost twice as likely to report excellent health (OR: 1·87, 95% CI: 1□15 to 3□08) and had, on average, a 0□59 (95%CI: 0□30 to 0□88) unit higher life satisfaction score than those not meeting any recommendations. The ORs for excellent health and the regression coefficients for life satisfaction increased relative to the number of recommendations met (Table 3). We also observed an association between adherence patterns of lifestyle behaviours during the second wave and well-being (Table 4). In addition to participants meeting the recommendations for all three lifestyle behaviours, participants meeting recommendations for PA and sleep, PA and ST, and sleep and ST were also more likely to report excellent health and had higher average life satisfaction scores than those not meeting any recommendation.

**Table 3.** Adjusted associations between number of recommendations met and well-being.

|  | Excellent health |  | Life satisfaction |  |
| --- | --- | --- | --- | --- |
| | aOR (95% CI) | p-value | $\beta$ (95% CI) | p-value |
| **Before pandemic** |  |  |  |  |
| No recommendation met | Reference |  | Reference |  |
| One recommendation met | 1.06 (0.62 to 1.83) | 0.834 | -0.17 (-0.49 to 0.15) | 0.308 |
| Two recommendations met | 1.24 (0.70 to 2.21) | 0.461 | -0.19 (-0.53 to 0.15) | 0.279 |
| All three met | 1.30 (0.71 to 2.39) | 0.400 | -0.05 (-0.41 to 0.32) | 0.802 |
| **During lockdown** |  |  |  |  |
| No recommendation met | Reference |  | Reference |  |
| One recommendation met | 0.89 (0.63 to 1.25) | 0.487 | 0.08 (-0.13 to 0.29) | 0.452 |
| Two recommendations met | 0.91 (0.62 to 1.33) | 0.626 | 0.13 (-0.11 to 0.36) | 0.293 |
| All three met | 1.15 (0.75 to 1.78) | 0.514 | 0.19 (-0.08 to 0.46) | 0.167 |
| **Second wave** |  |  |  |  |
| No recommendation met | Reference |  | Reference |  |
| One recommendation met | 1.34 (0.88 to 2.07) | 0.174 | 0.26 (0.01 to 0.51) | 0.038 |
| Two recommendations met | 1.57 (1.00 to 2.50) | 0.056 | 0.50 (0.23 to 0.77) | <0.001 |
| All three met | 1.87 (1.15 to 3.08) | 0.013 | 0.59 (0.30 to 0.88) | <0.001 |

Abbreviations: $\beta$ : regression coefficient; CI: Confidence interval; aOR: adjusted odds ratio.
Time points: before pandemic: before March 2020; during lockdown: between 16 March and 10 May 2020; second wave: between October 2020 and January 2021.
The three measures of number of recommendations met were included in a single model. Models were adjusted for child's sex, age category, BMI category, parental nationality, parental educational level, and region.

**Table 4.** Adjusted associations between recommendations adherence patterns and well-being.

|  | Excellent health |  | Life satisfaction |  |
| --- | --- | --- | --- | --- |
| | aOR (95%CI) | p-value | $\beta$ (95%CI) | p-value |
| **Before pandemic** |  |  |  |  |
| No recommendation met | Reference |  | Reference |  |
| PA only | 1.10 (0.53 to 2.28) | 0.801 | -0.04 (-0.48 to 0.39) | 0.841 |
| Sleep only | 1.10 (0.55 to 2.17) | 0.790 | 0.06 (-0.35 to 0.47) | 0.767 |
| ST only | 0.96 (0.53 to 1.76) | 0.905 | -0.33 (-0.69 to 0.03) | 0.069 |
| PA + Sleep | 1.43 (0.74 to 2.78) | 0.286 | -0.22 (-0.62 to 0.17) | 0.269 |
| PA + ST | 1.32 (0.68 to 2.58) | 0.413 | -0.13 (-0.53 to 0.27) | 0.529 |
| Sleep + ST | 1.18 (0.64 to 2.18) | 0.604 | -0.16 (-0.53 to 0.21) | 0.388 |
| All three | 1.30 (0.71 to 2.41) | 0.403 | -0.02 (-0.39 to 0.35) | 0.915 |
| **During lockdown** |  |  |  |  |
| No recommendation met | Reference |  | Reference |  |
| PA only | 1.11 (0.58 to 2.09) | 0.758 | 0.06 (-0.33 to 0.46) | 0.750 |
| Sleep only | 0.94 (0.53 to 1.66) | 0.823 | -0.02 (-0.38 to 0.34) | 0.907 |
| ST only | 0.86 (0.60 to 1.23) | 0.407 | 0.12 (-0.09 to 0.34) | 0.264 |
| PA + Sleep | 0.73 (0.34 to 1.51) | 0.404 | 0.26 (-0.19 to 0.72) | 0.259 |
| PA + ST | 1.06 (0.69 to 1.63) | 0.801 | 0.12 (-0.14 to 0.39) | 0.362 |
| Sleep + ST | 0.86 (0.57 to 1.31) | 0.489 | 0.15 (-0.11 to 0.41) | 0.247 |
| All three | 1.17 (0.75 to 1.81) | 0.489 | 0.22 (-0.05 to 0.49) | 0.112 |
| **Second wave** |  |  |  |  |
| No recommendation met | Reference |  | Reference |  |
| PA only | 1.02 (0.54 to 1.88) | 0.960 | 0.24 (-0.12 to 0.60) | 0.190 |
| Sleep only | 1.32 (0.79 to 2.23) | 0.290 | 0.25 (-0.06 to 0.55) | 0.119 |
| ST only | 1.50 (0.92 to 2.47) | 0.106 | 0.30 (0.01 to 0.60) | 0.042 |
| PA + Sleep | 1.86 (1.04 to 3.37) | 0.038 | 0.48 (0.12 to 0.84) | 0.009 |
| PA + ST | 1.48 (0.84 to 2.64) | 0.177 | 0.62 (0.27 to 0.96) | <0.001 |
| Sleep + ST | 1.59 (0.97 to 2.63) | 0.070 | 0.46 (0.16 to 0.75) | 0.003 |
| All three | 1.89 (1.15 to 3.15) | 0.013 | 0.57 (0.27 to 0.87) | <0.001 |

Abbreviations: $\beta$ : regression coefficient; CI: Confidence interval; aOR: adjusted odds ratio; PA: physical activity; sleep: sleep duration; ST: screen time.
Time points: before pandemic: before March 2020; during lockdown: between 16 March and 10 May 2020; second wave: between October 2020 and January 2021.
The three measures of adherence patterns were included in a single model. Models were adjusted for child's sex, age category, BMI category, parental nationality, parental educational level, and canton

Results for the models including lifestyle before the pandemic only and before the pandemic and during lockdown are presented in the appendix (Tables S9 and S10).

## DISCUSSION

In this population-based study, we found that the first two waves of the COVID-19 pandemic affected lifestyle behaviours in 5- to 18-year-olds in Switzerland. During the lockdown period between March and May 2020, the percentage of children and adolescents meeting the recommendations for PA and ST decreased drastically. Although it partially recovered during the second wave of the pandemic between October 2020 and January 2021, the percentage remained below the pre-pandemic period, especially for PA and for those aged 10 years or older. Changes in sleep duration were less marked and they were heterogeneous among age groups. Furthermore, we found an association between the number of recommendations met for lifestyle behaviours during the second wave and well-being assessed between January and April 2021. In contrast, there was no such association for the periods before the pandemic or during the lockdown. Participants meeting the recommendations for all three lifestyle behaviours or for combinations of two of them in the second wave were more likely to report excellent health and had a higher life satisfaction score, than those not meeting any recommendation.

Our findings of worsened lifestyle behaviours during the lockdown period are in line with previous cross-sectional and longitudinal studies in children and adolescents.^3^ In contrast to previous studies, we did not only analyse the early phases of the pandemic but extended our assessment until early 2021. This allowed us to show that, despite the reopening of schools and extracurricular activities for children and adolescents up to 16 years of age, children and adolescents continued to have a more sedentary lifestyle during the second wave of the pandemic compared to the pre-pandemic period, but partially recovered from the lockdown period. During the second wave, extracurricular activities such as sports training were partially open with strict restrictions on place including reduced times and numbers of participants, thus limiting the options to be physically active, especially indoors and during the winter months. In addition, due to the high incidence of cases during the second wave, parents may have felt less confident letting their children participate in group or indoor activities. Also, it is possible that the increase in screen-related activities resulted in the reduction of PA at least in part, as evidenced by the negative association between total ST and PA.^26^ The shift towards a more sedentary lifestyle during the pandemic, despite some recovery during the second wave, is of great concern for children’s and adolescents’ health and development. A cohort study of children in Austria reported that COVID-19 mitigation measures in 2020 were associated with an increase in the proportion of children with overweight and in a reduction of cardiorespiratory fitness,^27^ which may be partially explained by the reduction of PA and a concomitant increase in sedentary behaviour. Another study in school-aged children in China showed that home confinement during the pandemic was associated with an increase in the prevalence of myopia likely due to significantly decreased time spent outdoors and increased ST.^28^ It is also worrying that the pandemic exacerbated the sedentary lifestyle of older adolescents. Only a small percentage of adolescents aged 15-18 years, especially girls, met recommendations for PA and ST before the pandemic, and even a lower percentage met recommendations for all three lifestyle behaviours. Restrictions during the lockdown period resulted in only 22% and 5% of adolescents complying with PA and ST recommendations, respectively. The shift towards an even more sedentary lifestyle during the pandemic may place adolescents at an increased risk for deleterious long-term health effects such as cardio-metabolic risk and mental health problems.

We found that the pandemic also led to a change in adherence to recommendations for sleep in adolescents, but not in children aged 5-9 years. During the lockdown, the percentage of adolescents aged 10-14 and 15-18 years meeting the recommendations for sleep was slightly higher than before the pandemic, consistent with previous literature.^29^ In contrast, during the second wave, the percentage of compliant adolescents was lower than in the pre-pandemic period. This reduction may be explained by the increase in ST. Previous research reported that adolescents who spent more time on screens slept fewer hours, primarily due to time spent on portable devices such as phones, which proved to delay sleep onset.^30^ During the lockdown period, the irregular timetable and the absence of travel to school, may have allowed the delay in sleep time to be compensated by a later waking time, resulting in an overall increase in sleep duration. However, it is likely that this compensation may not have been possible during the second wave due to the return to a more stable schedule while schools were kept open.

We also showed that lifestyle is associated with two indicators of well-being, self-rated health and life satisfaction, which is consistent with previous research conducted before^8,9,18^ and during the pandemic.^10–12^ In contrast to previous studies, we accounted for the combined associations of PA, ST, and sleep duration with well-being, and demonstrated that meeting recommendations for all three behaviours as well as for combinations of two of them is more relevant for achieving optimal well-being than meeting recommendations for just a single behaviour. This supports previous literature showing a dose-response relationship between lifestyle behaviours and physical health or global cognition.^16,17^ Importantly, the possibility to include repeated measures of lifestyle allowed us to show that lifestyle behaviours during the second wave were predictors of well-being between January and April 2021, while there was no such association for the periods before the pandemic and during lockdown. This suggests that well-being in children and adolescents is associated with recent lifestyle behaviours and thus likely fluctuating as these behaviours change. To note, well-being was assessed in a separate follow-up questionnaire two to three months after the assessment of lifestyle behaviours during the second wave, thus ruling out potential bias attributable to cross-sectional designs.

Given the already high prevalence of children and adolescents not meeting lifestyle recommendations in the pre-pandemic period and the worsening during the pandemic in 2020, public health policies aiming to avoid permanent changes in lifestyle and negative consequences associated with adverse changes during the pandemic are urgently needed. Future research should test intervention strategies to revert lifestyle changes using specific approaches for different age groups, especially for adolescents. In addition, future research should assess the long-term consequences of the pandemic on lifestyle behaviours and extend the assessment to the period when all restrictions are lifted and, eventually, after the pandemic. Finally, our findings support the importance of taking an immediate holistic approach when designing public health strategies to promote lifestyle change and well-being.

The strengths of this study are the large sample size and the broad age range of the study sample, which allowed us to assess changes in lifestyle behaviours separately for different age groups. The population-based nature of the study and the inclusion of children and adolescents from four geographically and culturally different cantons of Switzerland support the external validity of our results. Finally, the availability of repeated measurements of different lifestyle behaviours allowed us to assess the associations between these behaviours alone and their combinations with reference to three different periods (i.e., pre-pandemic, lockdown, and second wave) and well-being measured in early 2021, almost one year after the onset of the pandemic.

We need to acknowledge some limitations. Lifestyle behaviours were self-reported or based on parental-report, with potential bias in the estimation of PA, ST, and sleep. In ZH, questions for PA referred to hours per day (instead of hours per week), which could have led to an overestimation of PA for this subgroup. Well-being indicators were also reported by parents for younger children and therefore some bias is possible for these measures. In addition, the fact that participants included in the study were more often from families with Swiss nationality and a high education level than those excluded due to missing data may not allow the generalizability of our results to populations with more ethnic variability and lower socio-economic status. Finally, although we accounted for a wide range of potential confounders when assessing the associations between lifestyle and well-being, residual confounding may still be a concern as we did not consider other factors such as existing psychological problems, parental mental health or family cohesion.

In conclusion, the COVID-19 pandemic has had a negative effect on children’s and adolescents’ lifestyle behaviours up to 2021, but some recovery has taken place. Policymakers should imperatively consider the balance of disease prevention and promotion of a healthy lifestyle when (re-)activating restrictive measures. In addition, future public health strategies aiming to promote children’s and adolescents’ well-being should target sufficient time for PA and sleep as well as reduce ST.

## Data Availability

Reasonable requests to share data will be considered by the Corona Immunitas steering committee subject to institutional agreements and ethics approvals. Data requests should be sent by email to the corresponding author.

## CONTRIBUTORS

SK, MAP, ALC, CK conceived the study. ALC, CK, AN, TR, AU, SK recruited study participants, collected and managed the data. SRH analysed the data. ALC, AN, GPP accessed and verified the data. GPP wrote the first draft. All authors contributed to the design of the study, the interpretation of results and revised and approved the manuscript for intellectual content. All authors confirm that they had full access to all the data in the study and accept responsibility to submit for publication.

## ACKNOWLEDGMENTS

We thank the Swiss Federal Statistical Office for providing the randomized list of participants. We thank SSPH+ for their help with coordination of all study sites. We also thank the ‘‘Volkschulamt” of the canton of Zurich for providing us with the comprehensive list of all schools and classes of the canton for the *Ciao Corona* study, and the supportive collaboration of school principals and teachers. In addition, we would like to thank all nurses, medical students, researchers, clinicians, and staff that contributed to the implementation of Corona Immunitas. Finally, we truly thank all children, adolescents, and their parents for their willingness to participate in this study.

