## appendix for "Lifestyle behaviours of children and adolescents during the first two waves of the COVID-19 pandemic in Switzerland and their relation to well-being: a population-based study"

**Description of study participants and data collection per canton**

This population-based observational study is part of *Corona Immunitas*, a research network that investigates the spread and impact of COVID-19 pandemic in Switzerland.[1] We included children and adolescents from four (out of 26) Swiss cantons: Ticino (TI), St. Gallen (SG), Graubünden (GR), and Zurich (ZH). These cantons belong to three out of four language regions (German, Italian and Romansch) in Switzerland and include 30% of the Swiss population.[2] Details on participants recruitment in each canton are presented below:

<u>TI:</u>

The data for Italian-speaking Switzerland stem from the *Corona Immunitas Ticino* population-based, prospective cohort study. It combines repeatedly collected digital survey and serology data from a representative sample during the course of the pandemic, with a specific focus on children and adolescents between 5 and 19 years of age. A representative sample of 4264 children and adolescents was drawn from the Swiss Federal Registry in May 2020 and invited to the study between September and November 2020. The study excluded people with insufficient Italian skills, under legal guardianship, in an asylum procedure, or with a short-term residency permit (i.e., < 1 year). Of all invited people, 1262 (30%) agreed to participate. Of these, 886 participants aged 5 to 18 years filled out baseline and follow-up questionnaires and provided data on the key variables for the present study. For children aged 5 to 13, parents provided answers with reference to the invited child. For adolescents aged 14 to 19, survey responses were based on self-report.

The study was approved by the Ethics Committee of the Canton of Ticino, Switzerland (2020-1514).

<u>SG and GR:</u>

The data for Eastern-Switzerland was collected over the course of the *Corona Immunitas Ostschweiz* in the cantons of St. Gallen and Graubünden. The study contains a onetime serology and digital survey data with monthly and weekly digital follow-up questionnaires. Based on a randomized sample drawn from the Swiss Federal Registry in October 2020, a total of 3335 children and adolescents aged 5 to 19 years were invited to participate between December 2020 and April 2021. Of all invited children, 523 (16%) agreed to participated. Of these, 395 participants aged 5 to 18 years filled out baseline and follow-up questionnaires and provided data on the key variables for the present study. For children aged 5 to 13, parents provided answers with reference to the invited child. For adolescents aged 14 to 19, survey responses were based on self-report.

The study was approved by the Ethics Committee of the Cantons of St. Gallen and Graubünden, Switzerland (2020-01247).

<u>ZH:</u>

The data for the Zurich canton come from the *Ciao Corona* study, which has been described elsewhere.[3] Briefly, *Ciao Corona* is a longitudinal, population-based observational study in a regionally representative cohort of children and adolescents (aged 6 to 16 years) attending school in the canton of Zurich, Switzerland. The study combines repeatedly collected serological and survey data. Primary schools were randomly selected from the list of all schools in the canton of Zurich, stratified by region, and matched with the geographically closest secondary school. Main exclusion criteria for schools were small school size (<40 children in a selected school level), and for participants—suspected or confirmed infection with SARS-CoV-2 during testing. Of 156 schools invited to participate, 55 schools agreed and 2585 children and adolescents were enrolled from June 16 to July 9, 2020. Of these, 1887 participants filled out baseline and follow-up questionnaires and provided data on the key variables for the present study. Parents/legal guardians of the participating children were invited to fill baseline and follow-up questionnaires online, together with their child.

The study was approved by the Ethics Committee of the Canton of Zurich, Switzerland (2020-01336).

**Additional details on statistical analysis**

We conducted descriptive analysis stratified by age category. We calculated 95% confidence intervals for percentages using the Wilson method.[4] To assess the associations of the number of recommendations met and the adherence patterns with overall health and life satisfaction, we used multivariable logistic and linear regression, respectively. We adjusted the models for child's sex, age category, BMI category, parents' nationality, educational level, and canton. We first tested a model including only the number of recommendations met/adherence patterns before the pandemic and the covariates. Then, we added the lifestyle measures for the lockdown period and, finally, for the second wave. In the model including the three time points, we tested interactions between lifestyle behaviours at the second wave and age category. However, the Akaike information criterion (AIC) and the p-value for the interactions indicated that models without interactions fit the data best:

| **Outcome-Exposure** | **p-value interaction** | **AIC model without interaction** | **AIC model with interaction** |
|---|---|---|---|
| Excellent health - Number of recommendations met | 0.098 | 2840 | 2842 |
| Excellent health - Adherence patterns | 0.388 | 2858 | 2871 |
| Life satisfaction - Number of recommendations met | 0.207 | 7120 | 7124 |
| Life satisfaction - Adherence patterns | 0.187 | 7136 | 7145 |

We used a complete case strategy for the multivariate analysis and reported missing data in the Table 1 footnotes. A further analysis, imputing well-being indicators and covariates yielded very similar results (data not shown). We performed the statistical analysis in R (version 4.0.3)[5] and used *ggplot2*[6] and *patchwork*[7] packages to produce graphs.

**Table S1. Details on data collection per canton†**

| | | ZH | TI* | SG/GR* |
|---|---|---|---|---|
| **Assessment** | | | | |
| **Baseline** | Date completed | June to October 2020 | October to December 2020 | December 2020 to January 2021 |
| | Reference for lifestyle questions | • Before pandemic<br>• During lockdown (March to May 2020) | • Before pandemic<br>• During lockdown (March to May 2020)<br>• Current moment of data collection | • Before pandemic<br>• During lockdown (March to May 2020)<br>• Current moment of data collection |
| **First follow-up** | Date completed | January 2021 | January to February 2021 | March to April 2021 |
| | Reference for lifestyle questions | • Current moment of data collection | NA | NA |
| | Reference for well-being questions | NA | • Current moment of data collection | • Current moment of data collection |
| **Second follow-up** | Date completed | March to April 2021 | NA | NA |
| | Reference for well-being questions | • Current moment of data collection | NA | NA |

Abbreviations: GR: Graubünden: SG: St. Gallen; TI: Ticino; ZH: Zurich; NA: Not Available

† In all cantons, baseline and follow-up questionnaires were open for a longer period than the period considered in the present analysis. However, we selected the period that corresponded to the second wave of the pandemic for lifestyle behaviours, and the closest periods between cantons for well-being measures.

* In TI, SG and GR, participants completed questionnaires following a rolling enrolment. Therefore, participants received the follow-up questionnaire three months after completing the baseline questionnaire.

**Table S2. Question formulation for lifestyle behaviours per canton**

| **Lifestyle behaviour*** | **ZH** | **TI** | **SG/GR** |
|---|---|---|---|
| Physical activity | How many hours of sport or other physical activity per day (that caused sweating or breathing more) does your child do on a typical weekday (counting school sport)?<br><br>How many hours of sport or other physical activity per day (that caused sweating or breathing more) per day does your child do on a typical weekend day (counting school sport)? | On average, during a week, how much time does your child spend with physical activity (at least with light sweating)? | On average, how much time does your child spend being physically active (with at least light sweating) during the week? (Including physical education at school) |
| Sleep | How many hours per day does your child sleep on a typical weekday? | On average, during a normal weekday (Mon-Fri), how many hours does your child sleep?<br><br>On average, during a normal weekend day (e.g., Sunday), how many hours does your child sleep? | On average, how many hours does your child sleep on a normal weekday (Mon-Fri)?<br><br>On average, how many hours does your child sleep on a normal holiday (e.g., Sunday)? |
| Screen time | How many hours a day does your child use electronic devices (e.g., mobile phone, play-station, Xbox, Nintendo, computer, TV)? | On average, during a normal weekday (Mon-Fri), how many hours does your child spend with electronic media (e.g., smartphone, computer, PlayStation, Xbox, Nintendo, TV?<br><br>On average, during a normal weekend day (e.g., Sunday), how many hours does your child spend with electronic media (e.g., smartphone, computer, PlayStation, Xbox, Nintendo, TV? | On average, how many hours does your child use electronic devices (e.g., smartphone, computer, Playstation, Xbox, Nintendo, TV) on a normal weekday (Mon-Fri)?<br><br>On average, how many hours does your child use electronic devices (e.g., smartphone, computer, Playstation, Xbox, Nintendo, TV) on a normal holiday (e.g., Sunday)? |

Abbreviations: GR: Graubünden: SG: St, Gallen; TI: Ticino; ZH: Zurich

* When asked for weekdays and weekend days separately, we calculated a weighted average as follows: [(weekday*5) + (weekend day*2)]/7.

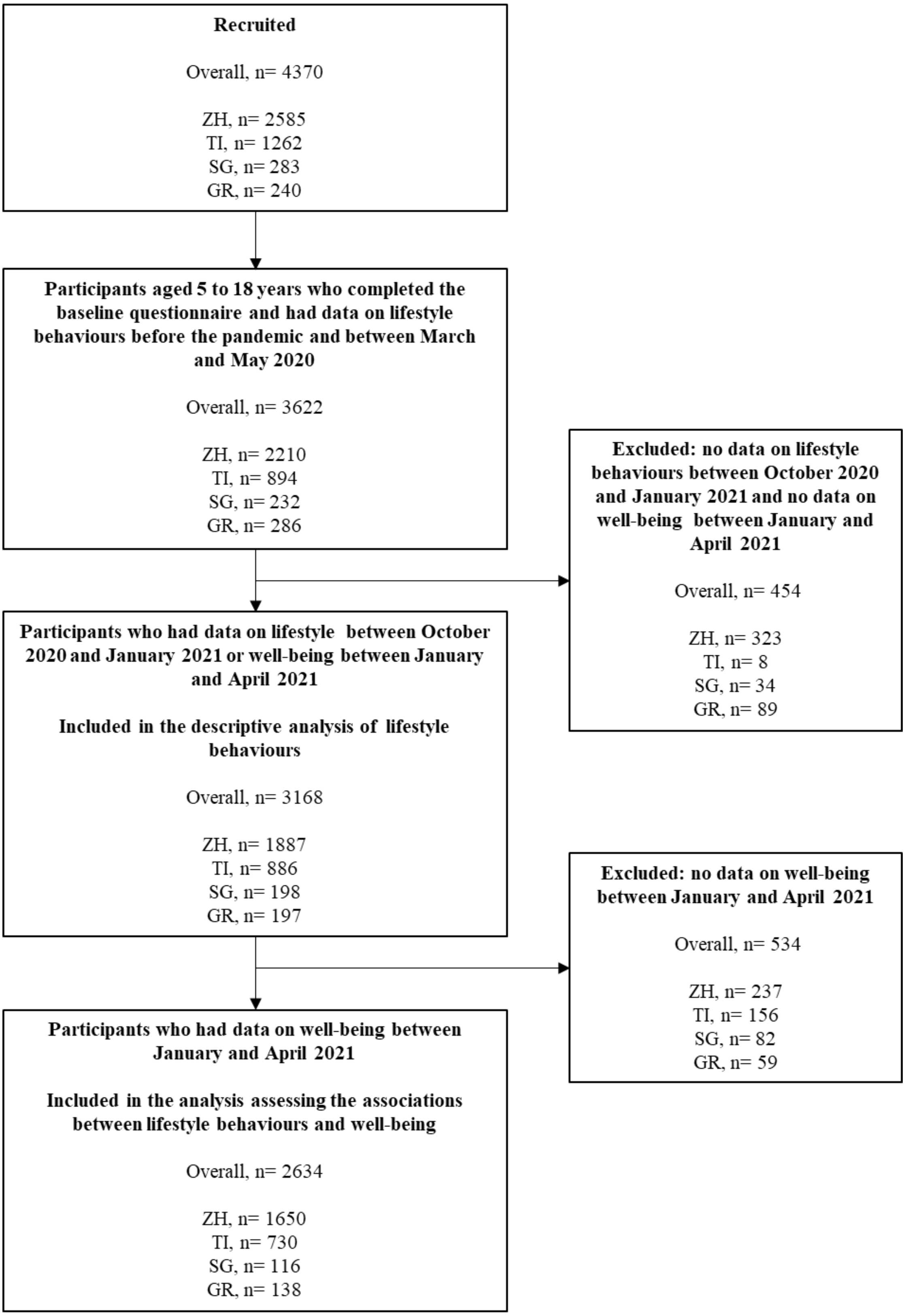

**Figure S1. Flowchart of the study population**

Abbreviations: GR: Graubünden: SG: St. Gallen; TI: Ticino; ZH: Zurich

**Table S3. Characteristics of children and adolescent included and excluded from the descriptive analysis**

| Characteristics | | Included<br>n= 3168 | Excluded*<br>n= 454 | p-value |
|---|---|---|---|---|
| **Sex. Girls** | | 1651 (52·1) | 235 (51·8) | 0·150 |
| **Age** | | | | 0·003 |
| | 5-9 | 1006 (31·8) | 113 (24·9) | |
| | 10-14 | 1734 (54·7) | 286 (63·0) | |
| | 15-18 | 428 (13·5) | 55 (12·1) | |
| **BMI category** | | | | 0·047 |
| | Underweight | 138 (4·4) | 20 (4·5) | |
| | Normal weight | 2487 (79·7) | 334 (75·1) | |
| | Overweight | 494 (15·8) | 91 (20·4) | |
| **Parental nationality** | | | | <0·001 |
| | Swiss | 2724 (86·3) | 326 (72·4) | |
| | Non-Swiss | 434 (13·7) | 124 (27·6) | |
| **Parental education** | | | | <0·001 |
| | Low/Medium | 837(27·1) | 155 (35·6) | |
| | High | 2247(72·9) | 280 (64·4) | |

Abbreviations: BMI: body mass index.

***** Excluded: participants who had no data on lifestyle between October 2020 and January 2021 nor well-being between January and April 2021.

Data are n (%). Some percentages do not add up to 100% because of rounding.

Some data were missing for the participants included (49 for BMI, 10 for parental nationality, and 84 for parental education) and excluded (9 for BMI, 4 for parental nationality, and 19 for parental education) from the analysis.

**Table S4. Characteristics of children and adolescent included and excluded from the analysis assessing the associations between lifestyle behaviours and well-being**

| Characteristics | Included<br>n= 2634 | Excluded*<br>n= 534 | p-value |
|---|---|---|---|
| **Sex. Girls** | 1369 (52·0) | 282 (52·8) | 0·699 |
| **Age** | | | |
| 5-9 | 879 (33·4) | 127 (23·8) | <0·001 |
| 10-14 | 1442 (54·8) | 292 (54·7) | |
| 15-18 | 313 (11·9) | 115 (21·5) | |
| **BMI category** | | | |
| Underweight | 116 (4·5) | 22 (4·2) | 0·306 |
| Normal weight | 2078 (80·1) | 409 (77·8) | |
| Overweight | 399 (15·4) | 95 (18·1) | |
| **Parental nationality** | | | |
| Swiss | 2303 (87·7) | 421 (79·1) | <0·001 |
| Non-Swiss | 323 (12·3) | 111 (20·9) | |
| **Parental education** | | | <0·001 |
| Low/Medium | 648 (25·2) | 198 (37·1) | |
| High | 1926 (74·8) | 321 (62·9) | |

Abbreviations: BMI: body mass index.

***** Excluded: participants who had no data on well-being between January and April 2021.

Data are n (%). Some percentages do not add up to 100% because of rounding

Some data were missing for the participants included (41 for BMI, 8 for parental nationality, and 60 for parental education) and excluded (8 for BMI, 2 for parental nationality, and 24 for parental education) from the analysis.

**Table S5. Prevalence of participants meeting recommendations for PA, ST and sleep by age group**

| | Age group | Before pandemic | During lockdown | Second wave |
|---|---|---|---|---|
| **PA** | **5-9 y** | 571/973 (58·7%) | 435/994 (43·8%) | 437/932 (46·9%) |
| | **10-14 y** | 1029/1634 (63·0%) | 673/1700 (39·6%) | 697/1594 (43·7%) |
| | **15-18 y** | 129/423 (30·5%) | 93/427 (21·8%) | 99/416 (23·8%) |
| **ST** | **5-9 y** | 929/993 (93·6%) | 753/992 (75·9%) | 865/931 (92·9%) |
| | **10-14 y** | 1250/1711 (73·1%) | 646/1693 (38·2%) | 1025/1596 (64·2%) |
| | **15-18 y** | 85/424 (20·0%) | 19/396 (4·8%) | 66/411 (16·1%) |
| **Sleep** | **5-9 y** | 901/998 (90·3%) | 865/997 (86·8%) | 847/937 (90·4%) |
| | **10-14 y** | 1248/1705 (73·2%) | 1358/1701 (79·8%) | 1058/1601 (66·1%) |
| | **15-18 y** | 263/424 (62·0%) | 293/423 (69·3%) | 231/416 (55·5%) |

Abbreviations: PA: physical activity; sleep: sleep duration; ST: screen time; y: years.

Time points: before pandemic: before March 2020; during lockdown: between 16 March and 10 May 2020; second wave: between October 2020 and January 2021.

Data are n/N (%). We classified participants as meeting recommendations according to international guidelines: ≥ 1h/day of PA; ≤2h/day of ST; 10-13 h/nigh of sleep for age 5 years, 9-11 h/night for ages 6-13 years, 8-10 h/night for ages 14-17, 7-9 h/night for age 18 years.

**Table S6. Number of recommendations met by age group**

| Age group | Time point | None | One | Two | All three |
|---|---|---|---|---|---|
| **5-9 y** | **Before pandemic** | 5 (0·5%) | 84 (8·8%) | 366 (38·3%) | 501 (52·4%) |
| | **During lockdown** | 31 (3·2%) | 190 (19·5%) | 439 (45·0%) | 316 (32·4%) |
| | **Second wave** | 6 (0·7%) | 89 (9·7%) | 446 (48·5%) | 379 (41·2%) |
| **10-14 y** | **Before pandemic** | 69 (4·3%) | 326 (20·5%) | 579 (36·4%) | 616 (38·7%) |
| | **During lockdown** | 174 (10·6%) | 628 (38·4%) | 557 (34·0%) | 277 (16·9%) |
| | **Second wave** | 148 (9·5%) | 468 (30·0%) | 572 (36·6%) | 374 (23·9%) |
| **15-18 y** | **Before pandemic** | 106 (25·5%) | 188 (45·3%) | 89 (21·4%) | 32 (7·7%) |
| | **During lockdown** | 98 (24·9%) | 219 (55·6%) | 69 (17·5%) | 8 (2·0%) |
| | **Second wave** | 125 (30·6%) | 192 (47·1%) | 76 (18·6%) | 15 (3·7%) |

Abbreviations: y: years.

Time points: before pandemic: before March 2020; during lockdown: between 16 March and 10 May 2020; second wave: between October 2020 and January 2021.

Data are n (%).

**Figure S2. Prevalence of participants meeting recommendations for PA, ST and sleep by age group and sex**

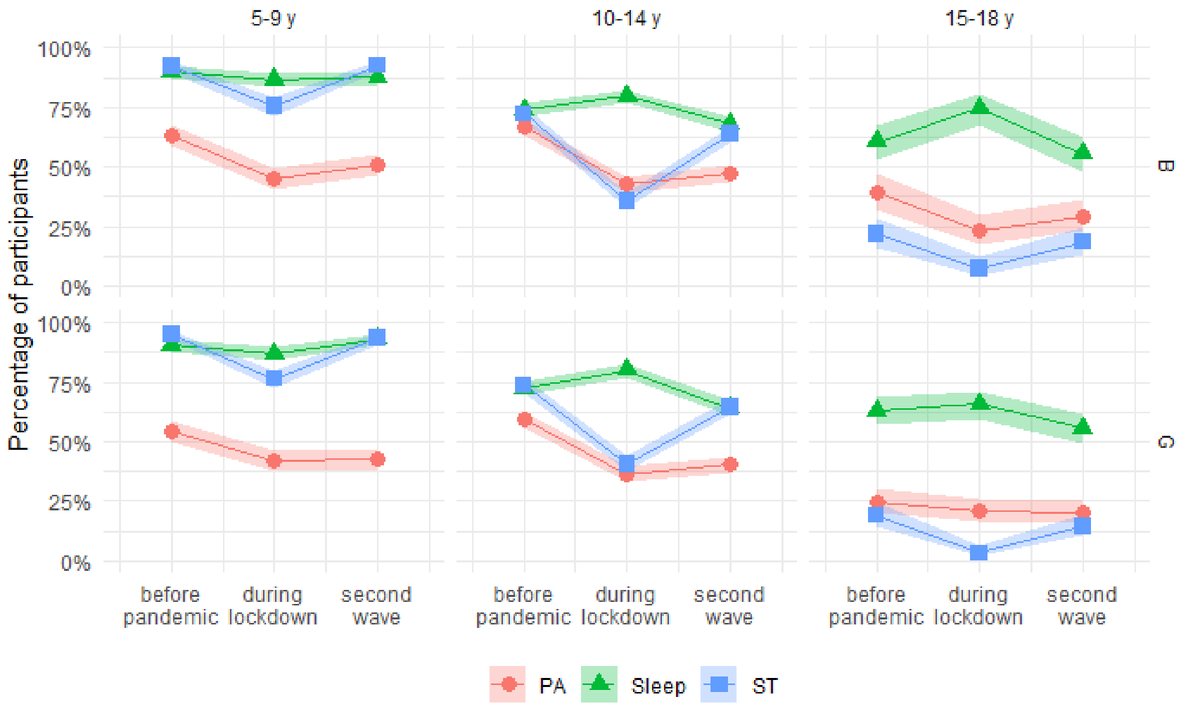

Abbreviations: B: boys; G: girls; PA: physical activity; sleep: sleep duration; ST: screen time.

Time points: before pandemic: before March 2020; during lockdown: between 16 March and 10 May 2020; second wave: between October 2020 and January 2021.

Shaded area represents 95% confidence intervals. We classified participants as meeting recommendations according to international guidelines: ≥ 1h/day of PA, ≤2h/day of ST, and the recommended range by age-groups for sleep duration (10-13 h/nigh for age 5 years, 9-11 h/night for ages 6-13 years, 8-10 h/night for ages 14-17, 7-9 h/night for age 18 years).

**Figure S3. Number of recommendations met period by age group and sex**

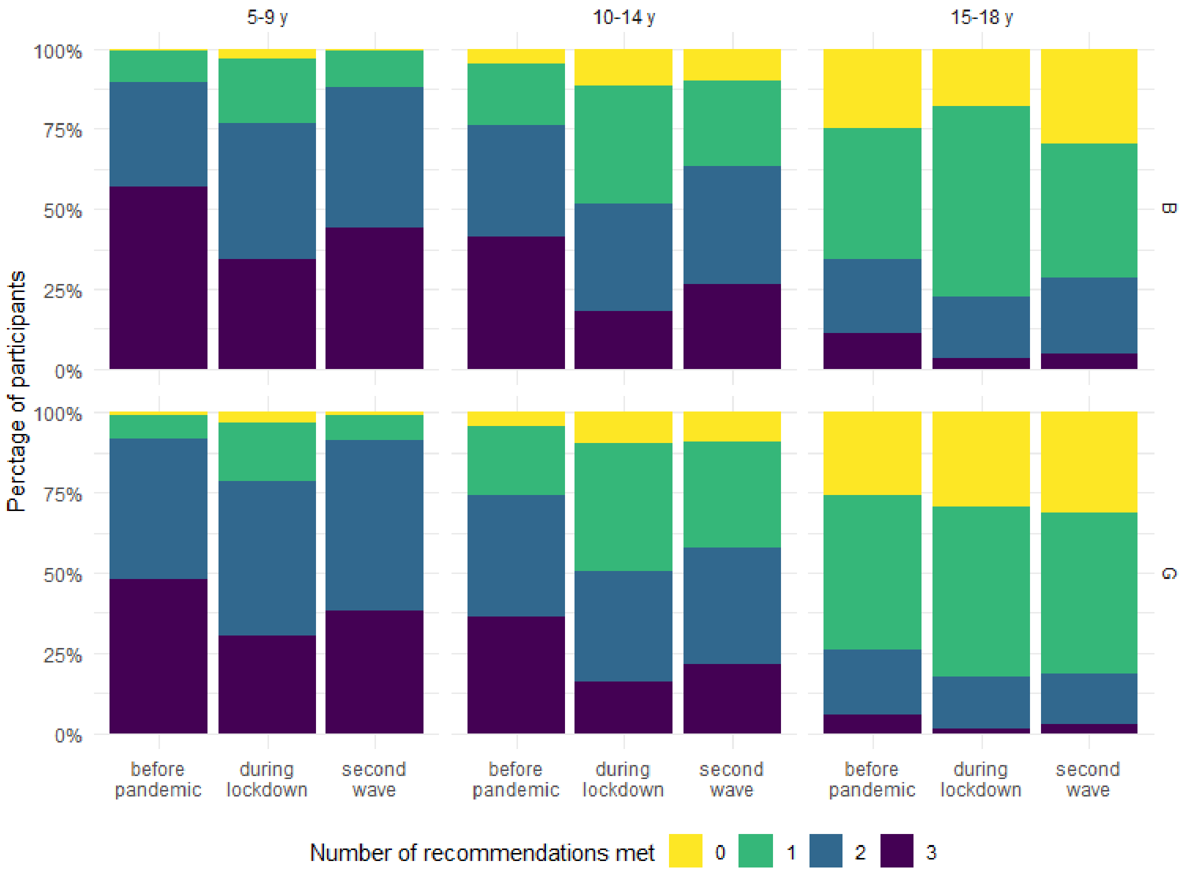

Abbreviations: B: boys; G: girls; y: years.

Time points: before pandemic: before March 2020; during lockdown: between 16 March and 10 May 2020; second wave: between October 2020 and January 2021.

**Table S7. Levels of PA, ST and sleep by age group**

| | | 5-9 y, n = 1,006 | 10-14 y, n = 1,734 | 15-18 y, n = 428 |
|---|---|---|---|---|
| **PA (h/day)** | **Before pandemic** | 1·0 (0·6 - 1·6) | 1·0 (0·7 - 1·9) | 0·6 (0·3 - 1·0) |
| | **During lockdown** | 0·6 (0·3 - 1·0) | 0·5 (0·3 - 1·0) | 0·4 (0·1 - 0·9) |
| | **Second wave** | 0·9 (0·6 - 1·4) | 0·9 (0·5 - 1·4) | 0·6 (0·3 - 0·9) |
| **ST (h/day)** | **Before pandemic** | 0·7 (0·5 - 1·0) | 1·5 (1·0 - 2·3) | 3·6 (2·5 - 5·0) |
| | **During lockdown** | 1·3 (0·8 - 2·0) | 3·0 (2·0 - 5·0) | 6·4 (4·9 - 8·2) |
| | **Second wave** | 1·0 (0·5 - 1·3) | 2·0 (1·0 - 3·0) | 3·7 (2·7 - 5·3) |
| **Sleep (h/night)** | **Before pandemic** | 10·0 (9·5 - 10·5) | 9·0 (8·0 - 9·6) | 8·0 (7·4 - 8·6) |
| | **During lockdown** | 10·0 (9·5 - 11·0) | 9·3 (9·0 - 10·0) | 8·9 (8·0 - 9·3) |
| | **Second wave** | 10·0 (9·3 - 10·3) | 9·0 (8·0 - 9·5) | 8·0 (7·3 - 8·4) |

Abbreviations: PA: Physical activity; sleep: sleep duration; ST: screen time, h: hours; y: years.

Time points: before pandemic: before March 2020; during lockdown: between 16 March and 10 May 2020; second wave: between October 2020 and January 2021.

Data are median ($P_{25}$-$P_{75}$).

**Table S8. Levels of PA, ST and sleep by age group and sex**

| | | Girls | | | Boys | | |
|---|---|---|---|---|---|---|---|
| | | **5-9 y, n = 514** | **10-14 y, n = 878** | **15-18 y, n = 259** | **5-9 y, n = 490** | **10-14 y, n = 855** | **15-18 y, n = 169** |
| **PA (h/day)** | **Before pandemic** | 1·0 (0·6 - 1·5) | 1·0 (0·6 - 1·6) | 0·6 (0·3 - 0·9) | 1·0 (0·7 - 1·8) | 1·1 (0·7 - 2·0) | 0·7 (0·4 - 1·4) |
| | **During lockdown** | 0·6 (0·3 - 1·0) | 0·5 (0·3 - 1·0) | 0·4 (0·1 - 0·7) | 0·7 (0·3 - 1·4) | 0·5 (0·3 - 1·0) | 0·3 (0·1 - 0·9) |
| | **Second wave** | 0·9 (0·6 - 1·2) | 0·9 (0·5 - 1·3) | 0·5 (0·3 - 0·9) | 1·0 (0·6 - 1·4) | 0·9 (0·5 - 1·5) | 0·6 (0·3 - 1·1) |
| **ST (h/day)** | **Before pandemic** | 0·6 (0·5 - 1·0) | 1·5 (1·0 - 2·3) | 3·6 (2·6 - 4·7) | 0·8 (0·5 - 1·1) | 1·5 (1·0 - 2·3) | 3·3 (2·3 - 5·0) |
| | **During lockdown** | 1·1 (0·7 - 2·0) | 3·0 (1·5 - 5·0) | 6·4 (5·0 - 8·0) | 1·3 (1·0 - 2·0) | 3·0 (2·0 - 5·0) | 6·4 (4·5 - 8·6) |
| | **Second wave** | 0·8 (0·5 - 1·3) | 2·0 (1·0 - 3·0) | 3·7 (2·9 - 5·1) | 1·0 (0·5 - 1·3) | 2·0 (1·0 - 3·0) | 3·7 (2·6 - 5·8) |
| **Sleep (h/night)** | **Before pandemic** | 10·0 (9·5 - 10·5) | 9·0 (8·0 - 9·6) | 8·0 (7·3 - 8·6) | 10·0 (9·0 - 10·3) | 9·0 (8·0 - 9·9) | 8·0 (7·6 - 8·5) |
| | **During lockdown** | 10·0 (10·0 - 11·0) | 9·5 (9·0 - 10·0) | 8·9 (8·0 - 9·3) | 10·0 (9·3 - 10·5) | 9·1 (9·0 - 10·0) | 8·6 (8·0 - 9·6) |
| | **Second wave** | 10·0 (9·5 - 10·3) | 9·0 (8·0 - 9·5) | 8·0 (7·3 - 8·4) | 10·0 (9·0 - 10·0) | 9·0 (8·0 - 9·5) | 8·0 (7·5 - 8·4) |

Abbreviations: PA: Physical activity; sleep: sleep duration; ST: screen time; h: hours; y: years

Time points: before pandemic: before March 2020; during lockdown: between 16 March and 10 May 2020; second wave: between October 2020 and January 2021.

Data are median ($P_{25}$-$P_{75}$).

**Table S9. Associations between number of recommendations met and well-being. Models including only measures before pandemic, and before pandemic and during lockdown.**

| | **Excellent health** | | **Life satisfaction** | |
|---|---|---|---|---|
| | **aOR (95% CI)** | **p-value** | **β (95% CI)** | **p-value** |
| **MODEL 1** | | | | |
| **Before pandemic** | | | | |
| No recommendation met | Reference | | Reference | |
| One recommendation met | 1·29 (0·83 to 2·02) | 0·256 | 0·27 (0·01 to 0·54) | 0·046 |
| Two recommendations met | 1·61 (1·04 to 2·52) | 0·035 | 0·42 (0·15 to 0·69) | 0·002 |
| All three met | 1·94 (1·23 to 3·10) | 0·005 | 0·69 (0·41 to 0·97) | <0·001 |
| **MODEL 2** | | | | |
| **Before pandemic** | | | | |
| No recommendation met | Reference | | Reference | |
| One recommendation met | 1·33 (0·84 to 2·14) | 0·224 | 0·25 (-0·03 to 0·53) | 0·085 |
| Two recommendations met | 1·63 (1·02 to 2·64) | 0·044 | 0·37 (0·08 to 0·66) | 0·011 |
| All three met | 1·78 (1·09 to 2·96) | 0·023 | 0·59 (0·29 to 0·90) | <0·001 |
| **During lockdown** | | | | |
| No recommendation met | Reference | | Reference | |
| One recommendation met | 1·02 (0·73 to 1·42) | 0·916 | 0·11 (-0·09 to 0·32) | 0·287 |
| Two recommendations met | 1·05 (0·74 to 1·51) | 0·782 | 0·16 (-0·06 to 0·39) | 0·154 |
| All three met | 1·35 (0·90 to 2·03) | 0·145 | 0·27 (0·01 to 0·52) | 0·039 |

Abbreviations: β: regression coefficient; CI: Confidence interval; aOR: adjusted odds ratio.
Time points: before pandemic: before March 2020; during lockdown: between 16 March and 10 May 2020; second wave: between October 2020 and January 2021.
Model 1 included only number of recommendations met before the pandemic, and model 2 included this measure for before the pandemic and during lockdown. Models were adjusted for child's sex, age category, BMI category, parental nationality, parental educational level, and canton.

**Table S10. Associations between recommendations adherence patterns and well-being. Models including only measures before pandemic, and before pandemic and during lockdown.**

| | Excellent health | | Life satisfaction | |
|---|---|---|---|---|
| | **aOR (95%CI)** | **p-value** | **β (95%CI)** | **p-value** |
| **MODEL 1** | | | | |
| **Before pandemic** | | | | |
| No recommendation met | Reference | | Reference | |
| PA only | 1·44 (0·78 to 2·65) | 0·241 | 0·38 (0·01 to 0·76) | 0·043 |
| Sleep only | 1·34 (0·77 to 2·36) | 0·305 | 0·43 (0·09 to 0·78) | 0·015 |
| ST only | 1·23 (0·77 to 1·99) | 0·387 | 0·16 (-0·13 to 0·45) | 0·275 |
| PA + Sleep | 1·82 (1·07 to 3·11) | 0·027 | 0·28 (-0·04 to 0·61) | 0·089 |
| PA + ST | 1·64 (0·97 to 2·79) | 0·068 | 0·51 (0·18 to 0·83) | 0·002 |
| Sleep + ST | 1·57 (0·99 to 2·5) | 0·057 | 0·46 (0·18 to 0·75) | 0·001 |
| All three | 2 (1·26 to 3·2) | 0·004 | 0·71 (0·43 to 0·99) | <0·001 |
| **MODEL 2** | | | | |
| **Before pandemic** | | | | |
| No recommendation met | Reference | | Reference | |
| PA only | 1·4 (0·73 to 2·68) | 0·311 | 0·34 (-0·05 to 0·74) | 0·088 |
| Sleep only | 1·33 (0·73 to 2·41) | 0·349 | 0·43 (0·07 to 0·79) | 0·021 |
| ST only | 1·28 (0·78 to 2·13) | 0·326 | 0·13 (-0·18 to 0·43) | 0·413 |
| PA + Sleep | 1·83 (1·04 to 3·24) | 0·037 | 0·24 (-0·10 to 0·59) | 0·169 |
| PA + ST | 1·65 (0·94 to 2·90) | 0·080 | 0·45 (0·11 to 0·80) | 0·010 |
| Sleep + ST | 1·58 (0·96 to 2·62) | 0·075 | 0·42 (0·11 to 0·72) | 0·007 |
| All three | 1·82 (1·11 to 3·02) | 0·020 | 0·62 (0·31 to 0·92) | <0·001 |
| **During lockdown** | | | | |
| No recommendation met | Reference | | Reference | |
| PA only | 1·31 (0·71 to 2·38) | 0·384 | 0·18 (-0·19 to 0·56) | 0·340 |
| Sleep only | 1·13 (0·65 to 1·95) | 0·664 | 0·05 (-0·3 to 0·39) | 0·791 |
| ST only | 0·99 (0·70 to 1·39) | 0·935 | 0·13 (-0·08 to 0·34) | 0·231 |
| PA + Sleep | 1·05 (0·53 to 2·05) | 0·891 | 0·37 (-0·06 to 0·79) | 0·094 |
| PA + ST | 1·15 (0·77 to 1·73) | 0·498 | 0·14 (-0·11 to 0·40) | 0·269 |
| Sleep + ST | 1·02 (0·69 to 1·52) | 0·907 | 0·16 (-0·08 to 0·41) | 0·193 |
| All three | 1·38 (0·92 to 2·08) | 0·123 | 0·28 (0·02 to 0·53) | 0·034 |

Abbreviations: β: regression coefficient; CI: Confidence interval; aOR: adjusted odds ratio; PA: physical activity; sleep: sleep duration; ST: screen time.

Time points: before pandemic: before March 2020; during lockdown: between 16 March and 10 May 2020; second wave: between October 2020 and January 2021.

Model 1 included only adherence patterns before the pandemic, and model 2 included this measure for before the pandemic and during lockdown. Models were adjusted for child's sex, age category, BMI category, parental nationality, parental educational level, and canton
